# Intraventricular Pressure Difference by Blood Speckle Tracking - Invasive Validation and Clinical Application

**DOI:** 10.1101/2024.08.20.24312326

**Authors:** Kristian Sørensen, Solveig Fadnes, Wadi Mawad, Matthew Henry, Hans Martin Flade, Andreas Østvik, Tor Åge Myklebust, Idar Kirkeby-Garstad, Lasse Løvstakken, Luc Mertens, Siri Ann Nyrnes

## Abstract

**Background:** Early diastolic relaxation creates an intraventricular pressure difference (IVPD) and resulting diastolic suction. Non-invasive estimation by echocardiographic techniques would allow to clinically evaluate this IVPD as an important component of ventricular filling. Recently, Blood Speckle Tracking (BST) echocardiography was introduced, allowing two-dimensional assessment of ventricular flow dynamics. Mitral inflow BST data can be used to estimate IVPD. The aims of the current study were to evaluate the accuracy of BST-based IVPD estimation compared to invasive pressure measurements in an in vivo animal model, and to clinically apply the method by comparing IVPD in children with univentricular hearts (UVH) and healthy controls.

**Methods:** The accuracy of BST-based IVPD-estimates was assessed in an open-chest porcine model, comparing BST-based IVPD with simultaneous repeated invasive pressure measurements in six pigs using micromanometer catheters. BST-based IVPD assessment was performed in 120 healthy controls and 44 patients with UVH < 18 years of age. Total IVPD (from base to apex) and apical IVPD (from the apical 2/3 of the ventricle) during early diastolic filling of the systemic ventricle was compared between patients with UVH and healthy controls.

**Results:** The validation in pigs included 103 measurements, demonstrating a mean difference of - 0.01mmHg (p=0.33) and high correlation (r = 0.95, p-value < 0.001) between IVPD from BST (-1.31 ± 0.28 mmHg) and invasive measurements (-1.30 ± 0.31 mmHg). In the pediatric patients, age range 2 days-17.76 years, feasibility was 96% in controls and 88.6% in UVH patients. Total and apical IVPD were significantly higher in controls compared to UVH (-1.82 vs -0.88 mmHg and -0.63 vs -0.33 mmHg, p < 0.001).

Variability was low with intraclass correlation coefficients of 0.99/0.96 (interobserver) and 0.98/0.99 (intraobserver) for total and apical IVPD respectively.

**Conclusions:** BST echocardiography provides accurate estimation of early diastolic IVPD. When clinically applied in children, we found high feasibility and reproducibility. IVPD was significantly lower in children with UVH compared to controls suggesting lower diastolic suction which can impact overall filling dynamics.

**Clinical perspective:** *What is new:*

- Blood speckle tracking echocardiography provides non-invasive estimation of intraventricular pressure difference in early diastole using two-dimensional blood flow velocities
- Intraventricular pressure difference based on blood speckle tracking is highly feasible, accurate and reproducible
- Blood speckle tracking demonstrates significantly reduced intraventricular pressure difference in early diastole in children with univentricular hearts indicating impaired relaxation and suction in these patients

*What are the clinical implications:*

- Intraventricular pressure difference based on blood speckle tracking is a novel and potential sensitive echocardiographic parameter to describe early diastolic ventricular relaxation and diastolic function in children with univentricular hearts
- Blood speckle tracking could improve assessment of diastolic function in children with congenital heart disease
- Non-invasive estimation of intraventricular pressure difference based on blood speckle tracking could improve assessment of diastolic function both in children and adults with heart disease

## Introduction

Congenital heart disease (CHD) is present in about 0.9% of live births and constitutes 28% of all major congenital anomalies.^1^ Advances in surgical and medical treatment have resulted in excellent outcomes with currently > 97% of children with CHD surviving into adulthood.

However, CHD still significantly impacts cardiovascular health throughout the lifespan with long-term risks of heart failure, arrhythmia, and stroke. This highlights the need for lifelong follow-up and monitoring of cardiovascular health.^2,3^ A key determinant of short and long term survival is the effect of CHD on cardiac function. While parameters for systolic function have been well established and are clinically used, non-invasive diastolic assessment is problematic in both children and adults with CHD. Especially in children, current diastolic echocardiographic parameters are incapable of reliably detecting diastolic dysfunction and filling pressures.^4^ This is related to the effect of age, heart rate and abnormal loading conditions on conventionally used diastolic parameters. Patients with CHD are at risk for developing diastolic dysfunction which significantly impacts their outcome, and therefore the development of novel techniques for assessing diastolic function is clinically important.^5,6^

Assessment of early relaxation is a key component in the evaluation of diastolic function.^7^ Early ventricular relaxation typically occurs earlier at the apex compared to the base of the heart which creates an intraventricular pressure difference (IVPD) and suction of blood from base to apex during early diastolic filling. ^8^ A comprehensive description of diastolic function should include assessment of diastolic suction.^9^ Therefore, an echocardiographic method to assess diastolic suction and IVPD could contribute to diastolic assessment.^10, 11, 12^

By combining high frame rate echocardiography with speckle-tracking technology, Blood Speckle Tracking (BST) enables visualization and estimation of two-dimensional (2D) intraventricular blood flow velocities without angle dependency and aliasing artefacts.^13^ BST can visualize complex flow patterns and provide quantitative flow data.^14, 15, 16, 17, 18^ Our group recently developed a method for calculating early-diastolic IVPD based on BST.^19^ The aim of the current study was to 1) validate BST-based IPVD assessment against invasive pressure measurements, 2) technically optimize the method for clinical use by studying the effects of frame rate (FR) and respiration on the IVPD measurements, and 3) apply BST-based IVPD assessment in children with univentricular hearts (UVH). We selected this population as diastolic dysfunction is prevalent in patients with UVH ^6, 20, 21^ and an important factor in the failing Fontan physiology affecting long-term outcome.^22, 23^

### Invasive validation of BST based IVPD estimates in a porcine model

The study was approved by The Norwegian Food Safety Authority (FOTS ID 20832). Six Noroc pigs (Hybrid of Dutch Z line and Norwegian landrace, TN 70, weight 38.8 ± 4.4 kg, three female) were studied in an open chest model. All pigs received standard care in compliance with the European Convention on animal care. The pigs received intramuscular ketamine 10 mg/kg and Metedominine 0.08 mg/kg, intravenous access was established, and complete anesthesia obtained by fentanyl 5-8 mcg/kg, thiopentone 3-4 mg/kg, ketamine 5-8 mg/kg and atropine 1 mg before intubation and mechanical ventilation. Amiodarone 4 mg/kg was administered after induction to improve hemodynamic stability. Anesthesia was maintained by continuous infusion of propofol 8 mg/kg/h, dexmedetomidine 4 mcg/kg/h and ketamine 5 mg/kg/h, and the animals were closely monitored according to the standard anesthetic protocol of the lab. A bolus of Ringer lactate 10 ml/kg followed by continuous infusion of Ringer lactate12 ml/kg/h and Dobutamine 1.5 - 3.5 mcg/kg/min was given to stabilize hemodynamics. Additional boluses of Ringer lactate were given as necessary. The animals were euthanized with pentobarbital 100 mg/kg after completion of the experiments.

After the thoracotomy, two micromanometer pressure catheters (Millar Instruments, Houston, Texas) were placed in the left ventricle (LV) by apical puncture. The catheters were calibrated in a body-tempered saline bath before placement. The basal catheter was advanced until echocardiography and pressure recording confirmed left atrial position, then pulled back until LV pressure was obtained. Basal positioning in the central LV inflow was confirmed with echocardiography. The apical catheter was advanced until LV pressure was obtained. Pacemaker-generated premature beats were regularly induced, and the subsequent prolonged diastasis utilized to equalize differences between the catheters caused by drift and hydrostatic pressure. Invasive pressure recordings and epicardial apical four-chamber (4CH) BST images were simultaneously acquired (Figure 1).

**Figure 1.**
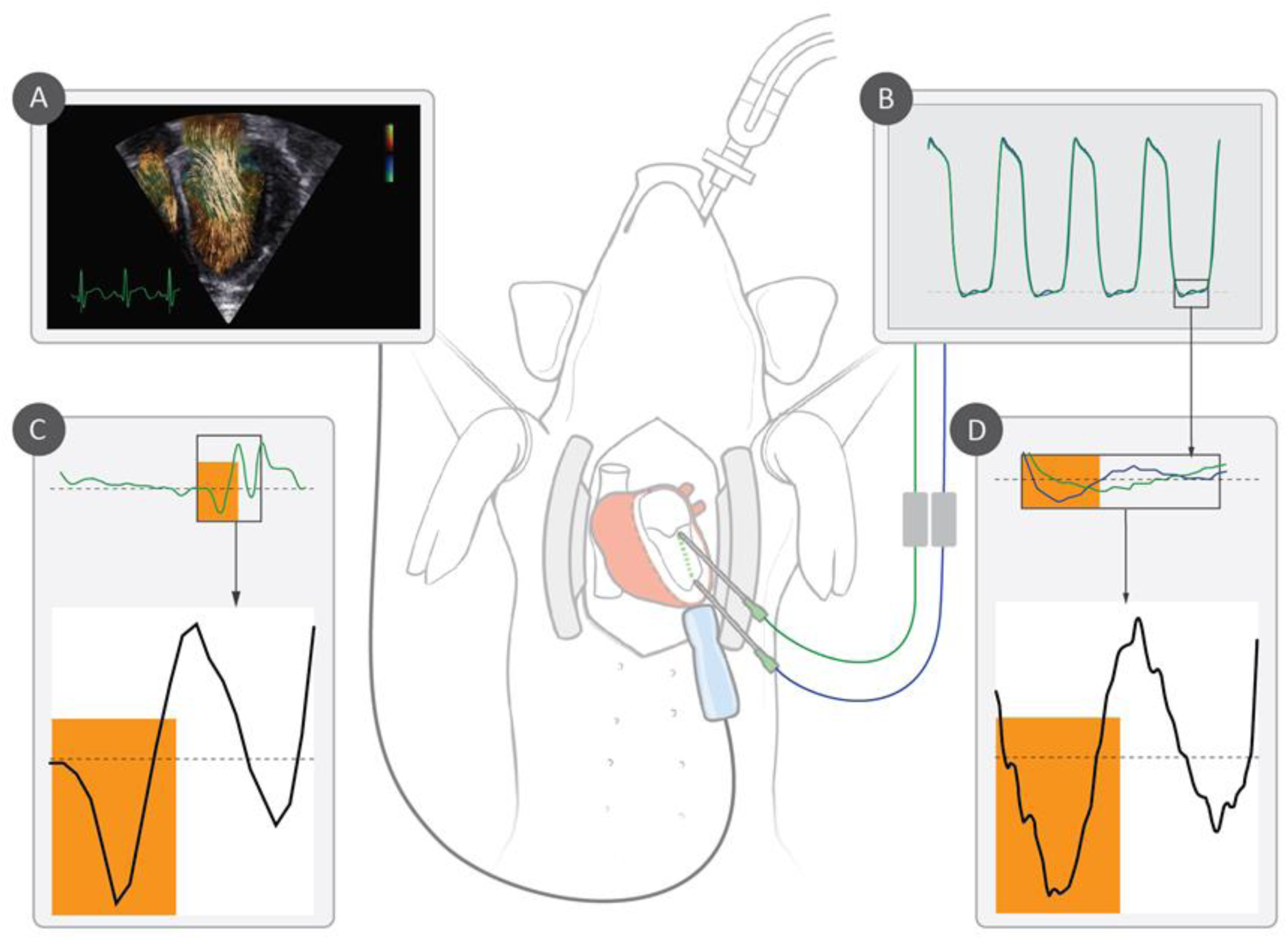
Invasive intraventricular pressure difference (IVPD) versus IVPD based on blood speckle tracking (BST) Apical left ventricular four chamber BST images (A) and invasive pressure recordings from two micromanometer catheters in the left ventricle (B) were simultaneously acquired. Invasive IVPD was calculated as the instantaneous pressure difference between the apical catheter (blue graph) and basal catheter (green graph) (B, D). IVPD in early diastolic filling from BST (C) was compared with the invasively measured IVPD (D).

Invasive IVPD was defined as the maximal instantaneous pressure difference between the basal and apical catheter during early diastolic filling. Invasive IVPD was compared to the IVPD estimate from BST from the same heartbeat. A synchronization signal ensured temporal matching of invasive and echocardiographic data. Only cycles with stable invasive pressure recordings, BST-acquisitions of sufficient quality and temporal matching were included in the analysis.

### Patient selection

Controls were included at Health Møre og Romsdal Hospital Trust in Ålesund, Norway.

Children < 18 years of age referred for cardiologic assessment due to palpitations, murmur or syncope were prospectively enrolled. To ensure controls across all ages, parents of healthy newborns were approached at the maternity ward and enrolled. Children with heart disease or chronic disease were not included. Patients < 18 years of age followed with UVH were prospectively enrolled at The Hospital for Sick Children, Toronto ON, Canada. The study was approved by The Regional Committee for Medical and Health Research Ethics, REC Central, and The Hospital for Sick Children Research Ethics Board. Written informed consent from parents or caregivers was obtained prior to enrollment.

### Equipment, image acquisition and processing

The participants underwent a clinical echocardiographic examination before the study images were obtained. Data from Health Møre og Romsdal Hospital trust were acquired by one pediatric cardiologist using a Vivid E90 ultrasound system (GE Healthcare, Norway) and the 6S phased-array probe. A second pediatric cardiologist acquired apical 4CH BST images in randomly selected controls for reproducibility analysis. Data from The Hospital for Sick Children were acquired by two pediatric cardiologists and two technicians using a Vivid E9/E95 ultrasound system (GE Healthcare, Norway) and the 6S phased-array probe.

All scanners had research software permitting high frame rate echocardiography-based BST acquisitions including storage of in-phase and quadrature data for offline processing.^13^ No sedative drugs were used during imaging for the study. EchoPAC version 202.50 and 204 and ViewPoint version 6.12.2 (GE Healthcare, Norway) were used for the clinical echocardiographic analysis.

### Total and regional IVPD estimation based on BST

BST acquisition based on high frame rate plane wave imaging and IVPD analysis based on BST were described previously.^13,19^ Flow data are acquired at FR equal to the Doppler pulse repetition frequency and combined with high-quality B-mode acquisitions in a duplex image setup. Blood flow velocity estimates are averaged for each duplex acquisition.

Apical 4CH BST acquisitions of systemic ventricular inflow in the mid-ventricular plane were exported to a custom-made software analysis program (PyUSView, NTNU, Norway) and analyzed by one pediatric cardiologist. B-mode of sufficient quality to allow identification of the anatomy of the systemic ventricle including the atrioventricular (AV) valve and apex and sufficient flow data to allow velocity estimation from base to apex in early diastole were required for the acquisition to be included in the analysis. Averaging in time (20 ms) and space (5x5 mm) was applied to reduce measurement variability. End systole was defined as the last frame before mitral valve (MV) opening. End diastole was defined as the first frame after MV closure.

BST acquisitions contain 2D velocity information from the entire flow field in every frame. In PyUSView, the pressure differences within the flow field are calculated from the velocity-changes in the flow field over time using the Navier-Stokes equations (supplemental material). A spline curve was drawn from the central part of the AV-orifice at the AV-junction, following the central body of LV inflow in the frame of early filling, to the apex of the ventricle. Total IVPD, defined as the maximal pressure difference between the AV-junction and apex along the spline curve in early filling, and apical IVPD, defined as the maximal pressure difference in the apical 2/3 of the ventricle along the spline curve during early filling, was automatically calculated by PyUSView (Figure 2).

**Figure 2.**
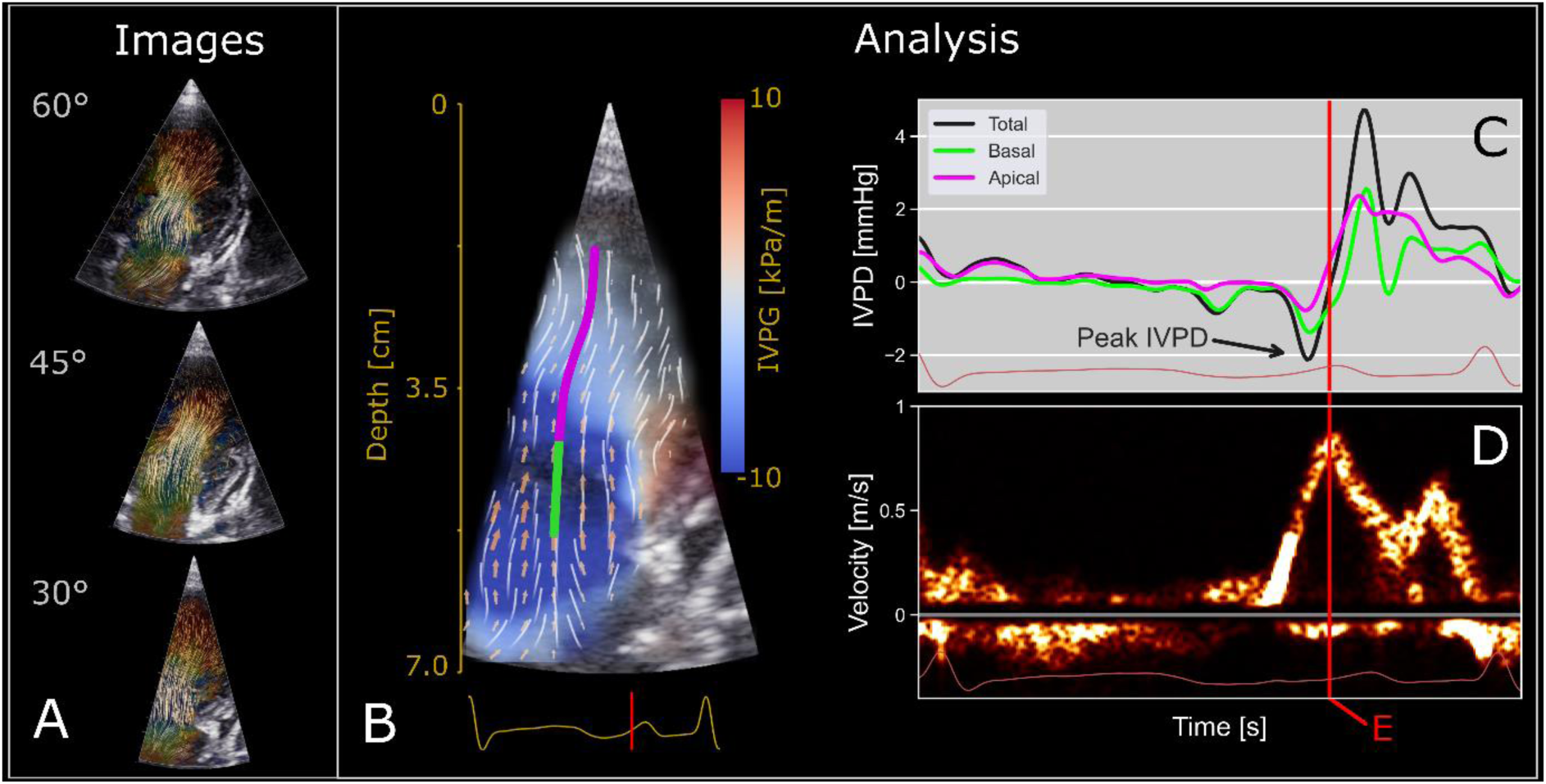
Blood speckle tracking acquisition and intraventricular pressure difference (IVPD) analysis. **(A)** Apical four-chamber acquisitions of left ventricular inflow in early diastole with different sector widths resulting in acquisitions with different frame rates. **(B)** Two-dimensional pressure difference map from IVPD analysis in PyUSView. Basal IVPD is calculated along the green part of the spline curve, apical IVPD is calculated along the purple part of the spline curve and total IVPD is calculated along the entire spline curve. **(C)** IVPD along the spline curve through the heart cycle. Peak total, basal and apical IVPD in early diastolic filling is automatically reported. **(D)** Peak IVPD precedes peak E-wave velocity (maximal inflow velocity of the systemic ventricle).

### Effect of FR on IVPD based on BST

Apical 4CH BST images with sector widths 60°, 45° and 30° were consecutively acquired in the controls (Figure 2A). Narrowing the image sector increases the FR. Keeping all other scanner-settings unchanged, the effect of FR on IVPD could be studied.

### Effect of respiration on IVPD based on BST

Due to the young age of many of the study participants, echocardiography was performed during spontaneous breathing. To investigate the effect of respiration on IVPD, multiple 4CH BST images with standardized scanner settings and registration of the respiratory curve were acquired in a subgroup of the controls.

### Reproducibility of IVPD based on BST

Reproducibility of postprocessing analysis of IVPD based on BST-acquisitions was previously demonstrated.^19^ To assess the dependence of IVPD reproducibility on image acquisition, two pediatric cardiologists acquired multiple apical 4CH BST images in 17 of the controls.

Postprocessing IVPD analysis from the acquisitions were performed by one observer with an interval of at least 4 weeks between analysis for inter- and intraobserver assessment respectively, and the results were compared.

### IVPD in UVH and controls

Total and apical IVPD were compared between controls and patients with UVH. IVPD was also compared between UVH with morphological right and morphological left ventricles and UVH patients at different surgical stages (bidirectional cavopulmonary connection (BCPC) and total cavopulmonary connection (TCPC) respectively).

### Statistical Analysis

Shapiro-Wilk test and visual inspection of histograms were used to assess the normality of continuous variables. Data are presented as median and interquartile range if nonnormally distributed and mean ± standard deviation if normally distributed. Categorical variables are presented as absolute and relative frequencies. Univariate tests included independent t-tests for difference in means and chi-square test for difference in distributions of categorical variables. Pearson correlation coefficient was used for estimating the strength of association between two variables. Agreement between IVPD from BST and invasive measurements was assessed using Bland-Altman plot and linear regression analysis. Inter- and intra-observer agreement was evaluated using the intraclass correlation coefficient (ICC) calculated with a two-way random effects model, treating raters as random effects. To incorporate the correlation introduced by repeated measures within patients, mixed models with random intercepts for each patient were used to compare IVPD measured at inspiration and expiration. P-values < 0.05 were considered statistically significant. Analyses were done using Stata version 18.0 and SPSS statistics version 29.

## Results

### Invasive validation of BST-based IVPD-estimates in a porcine model

A total of 103 heart cycles from the six pigs were eligible for analysis. Figure 1 illustrates that qualitatively the IVPD curves obtained by BST closely matched the corresponding invasive IVPD curves. Lower LV pressure is measured at the apex compared to the base (negative IVPD) during the first part of early diastolic filling followed by a reversed spatial pressure relationship (positive IVPD) in the last part of early diastolic filling.

Mean IVPD in early diastole was -1.30 ± 0.31 mmHg from the invasive measurements and -1.31 ± 0.28 mmHg from the non-invasive BST-estimation, giving a mean difference between the methods of -0.01 mmHg (p=0.33). Median Duplex FR of the BST acquisitions was 79 (68- 82). In Figure 3, a Bland-Altman plot comparing IVPD from the invasive measurements and BST shows no proportional bias and narrow 95% limits of agreement. There was a high linear correlation between IVPD from invasive measurements and BST (r = 0.95, p < 0.001).

**Figure 3.**
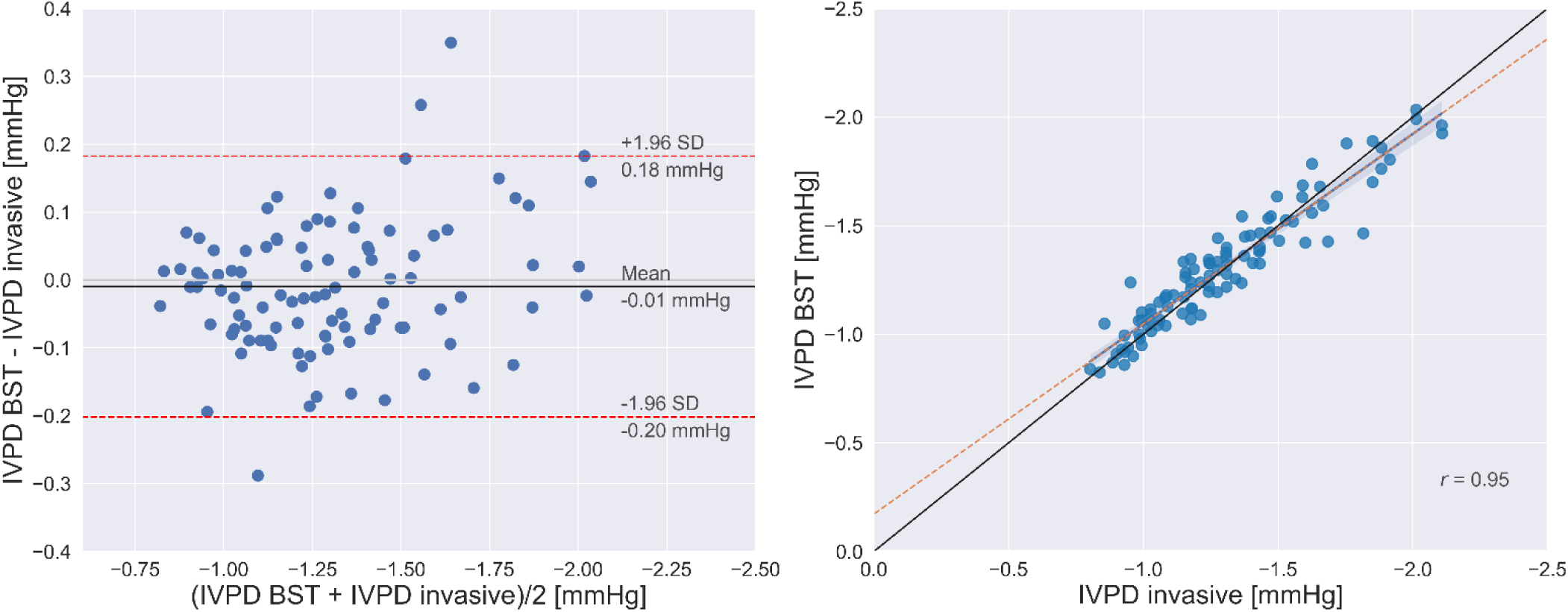
Comparisons between intraventricular pressure difference (IVPD) from blood speckle tracking (BST) and invasive measurements. Bland-Altman plot (left panel) shows a non-significant mean difference of 0.01 mmHg (p=0.33), no proportional bias and narrow 95% limits of agreement when comparing IVPD based on BST and invasive measurements. There was a high linear correlation between IVPD from invasive measurements and BST (r = 0.95, p < 0.001) (right panel).

### IVPD estimation based on BST

A total of 164 children, 120 controls and 44 UVH patients, were included in the study. Acquisition of three different sector widths in each control resulted in a total of 360 images in the 120 controls. After quality control, 118 controls (age range 2 days-17.76 years, 61 boys and 57 girls) had BST data eligible for IVPD analysis. From these, all 30° acquisitions were included while four 45° acquisitions and four 60° acquisitions were excluded due to insufficient image quality, giving a feasibility of 96% in the controls. Of the UVH patients (age range 7 months-15.25 years, 27 boys and 12 girls) 39/44 had BST data eligible for IVPD analysis, giving a feasibility of 88.6% in the UVH (Figure 4). Of the analyzed UVH patients, 20 had a dominant morphological right ventricle (RV) and 19 had a dominant morphological LV. Thirty-two were scanned after TCPC and 7 after BCPC.

**Figure 4.**
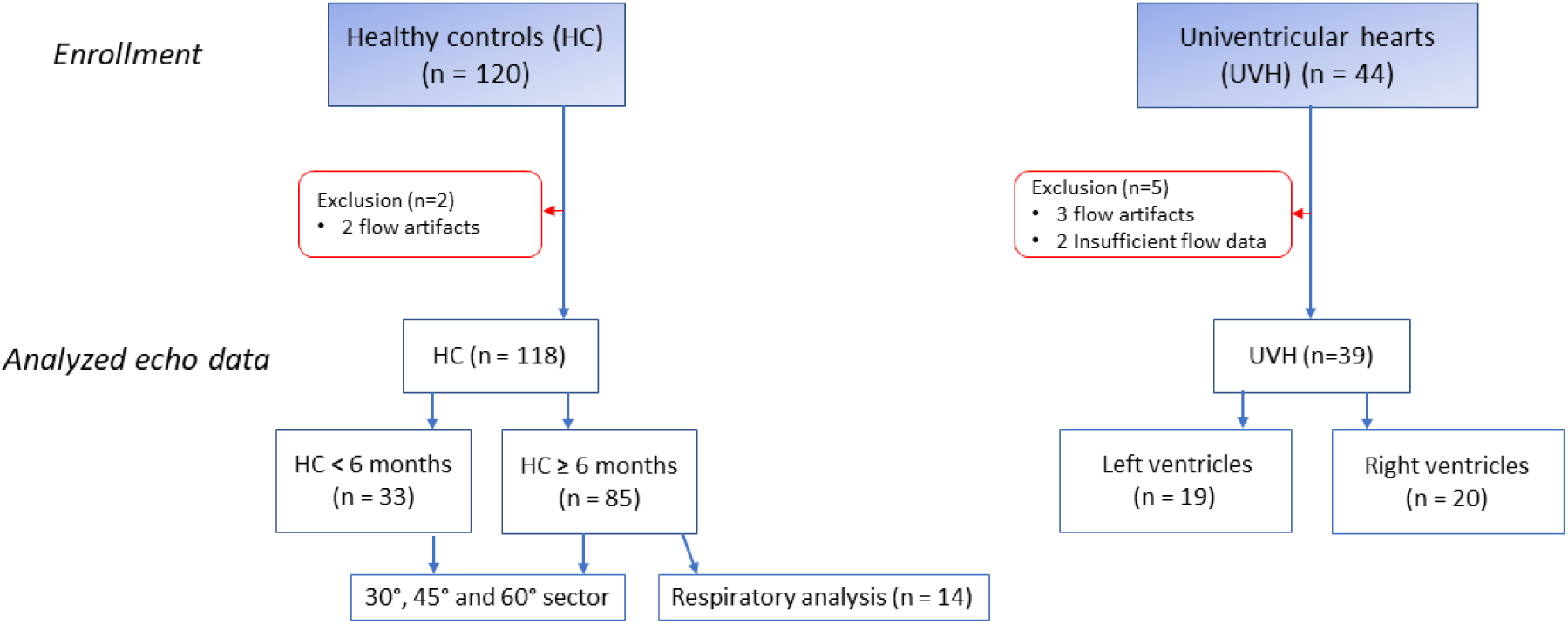
Subject enrollment.

### Effect of frame rate on IVPD based on BST

Narrowing of the sector width resulted in a significant increase in BST Duplex FR (sector width 60° median FR 45 (34.00-51.00), sector width 45° median FR 61 (47-68.75) and sector width 30° median FR 94 (76-107), P < 0.001 between all groups). Increased FR resulted in a statistically significant increase in both total and apical IVPD in the controls when comparing 60° with 45°, 45° with 30° and 60° with 30° sector, P < 0.000 between all groups (Table 1 and Figure 5C).

**Figure 5.**
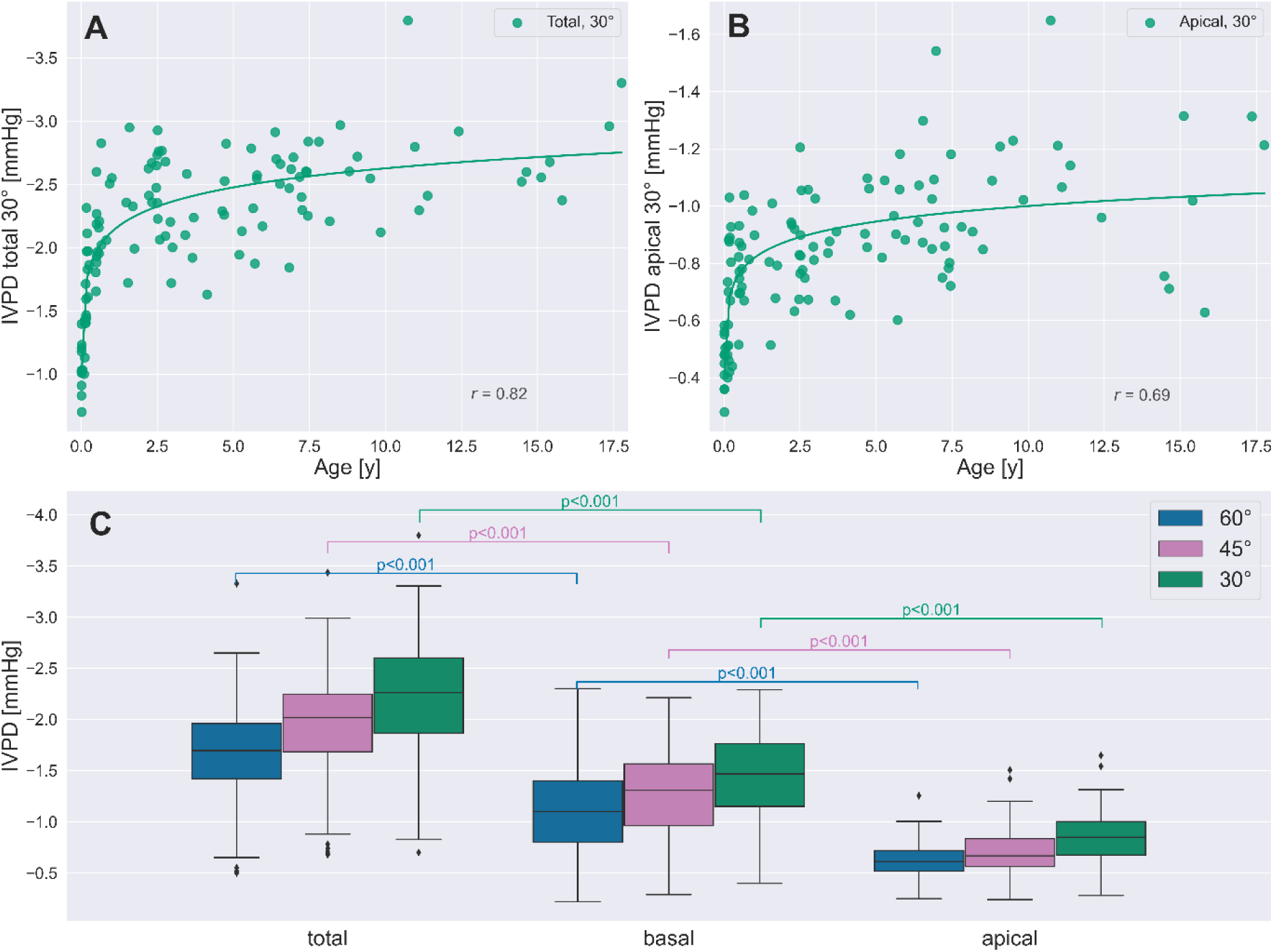
Intraventricular pressure difference (IVPD) in controls < 18 years of age. IVPD total **(A)** and IVPD apical **(B)** from acquisitions with 30° sector width versus age in healthy controls < 18 years of age. Box plot **(C)** comparing IVPD total, basal and apical from acquisitions with different sector widths in healthy controls. Narrowing of the sector width resulted in increased frame rates and a significant increase in total, basal and apical IVPD.

**Table 1.**
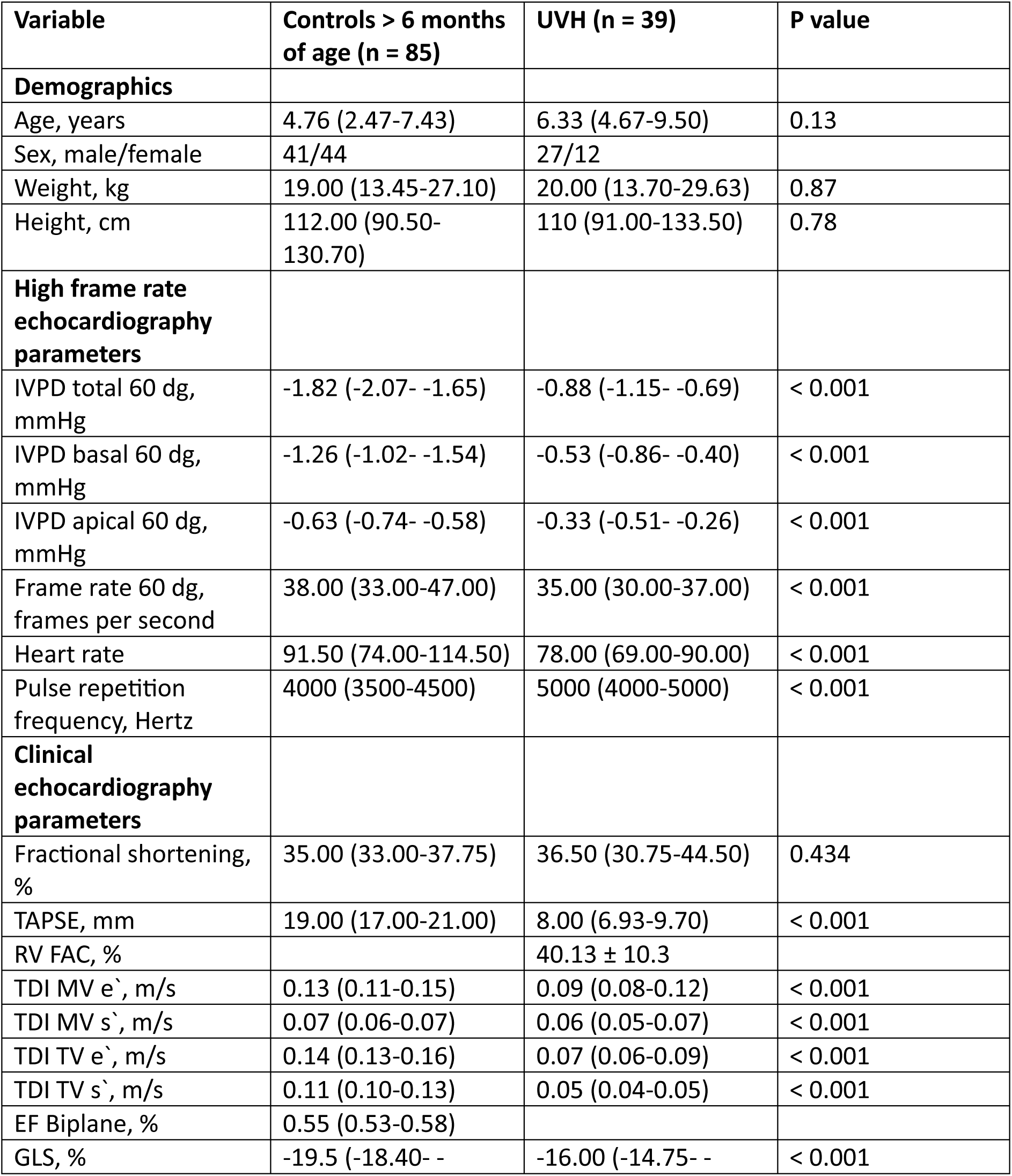
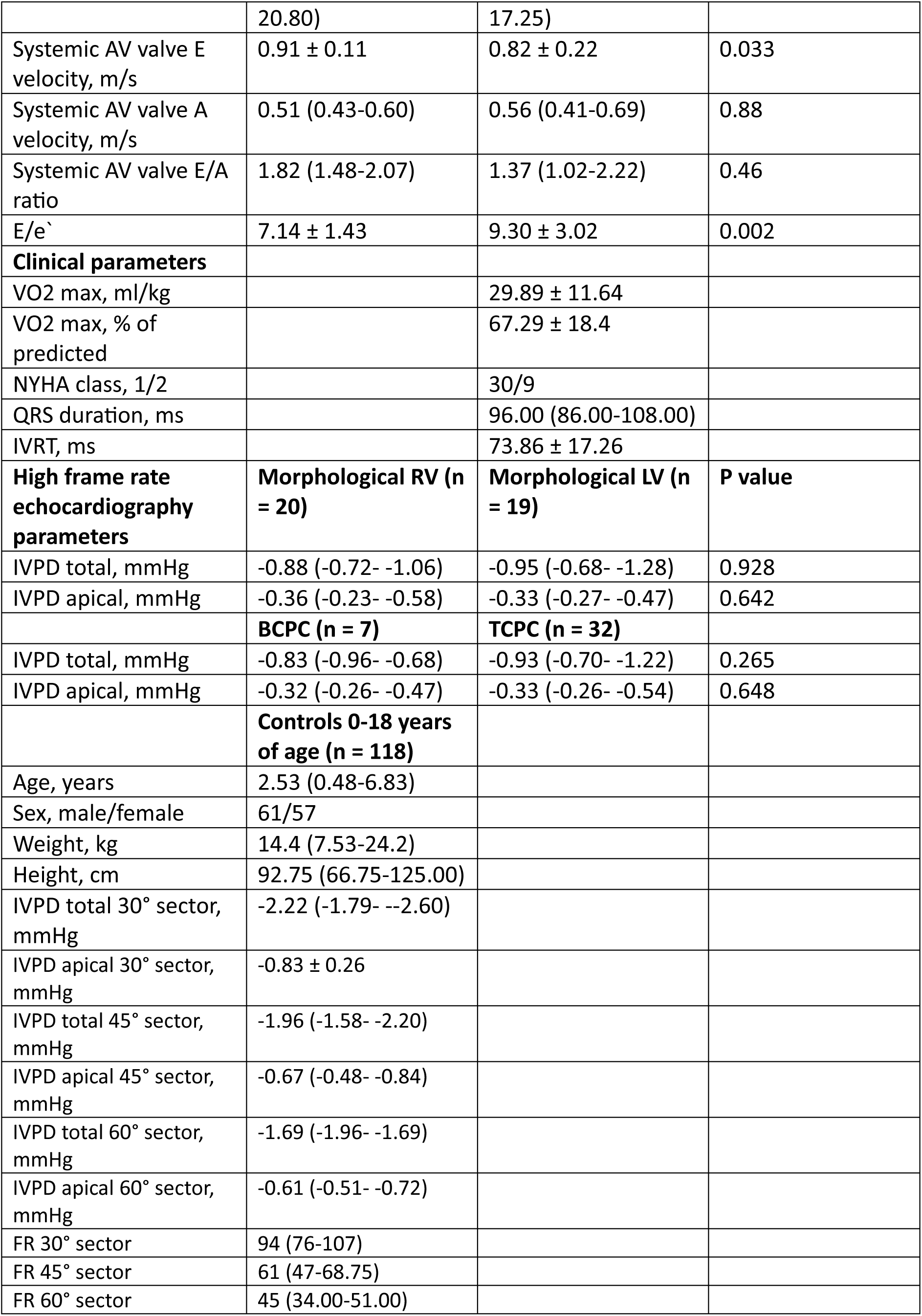
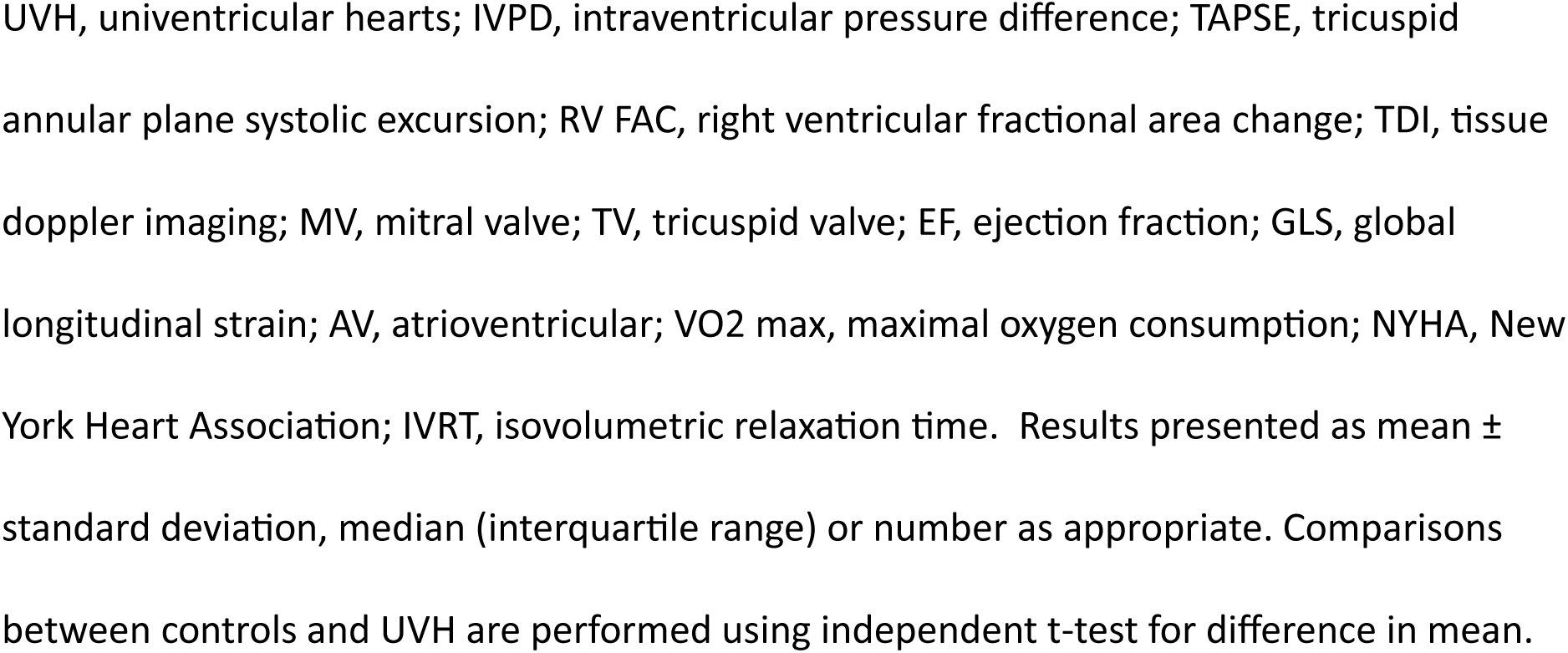
Clinical characteristics and echocardiographic data.

**Table 2.** Pearson correlation coefficients r between IVPD and demographic, echocardiographic and clinical parameters.

| Univentricular heart group |  |  |  |  | Control group > 6 months of age |  |  |  |
| --- | --- | --- | --- | --- | --- | --- | --- | --- |
| Variable | Correlation IVPD total |  | Correlation IVPD apical |  | Correlation IVPD total |  | Correlation IVPD apical |  |
|  | Pearson Correlation coefficient r | P value | Pearson Correlation coefficient r | P value | Pearson Correlation coefficient r | P value | Pearson Correlation coefficient r | P value |
| Age | -0.14 | 0.41 | 0.09 | 0.59 | -0.37 | 0.0006 | -0.10 | 0.37 |
| Fractional shortening, % | -0.13 | 0.76 | 0.26 | 0.53 | 0.21 | 0.055 | -0.12 | 0.29 |
| MAPSE, mm |  |  |  |  | -0.36 | 0.0008 | -0.21 | 0.0577 |
| TAPSE, mm | -0.38 | 0.17 | 0.25 | 0.38 | -0.35 | 0.001 | -0.12 | 0.27 |
| RV FAC, % | -0.04 | 0.91 | -0.10 | 0.78 |  |  |  |  |
| MV E velocity, m/s | -0.17 | 0.50 | -0.38 | 0.13 | -0.02 | 0.87 | -0.06 | 0.61 |
| MV A velocity, m/s | 0.18 | 0.47 | 0.19 | 0.46 | 0.22 | 0.046 | 0.15 | 0.18 |
| MV E/A ratio | -0.31 | 0.21 | -0.40 | 0.10 | -0.26 | 0.022 | -0.21 | 0.07 |
| TV E velocity, m/s | -0.23 | 0.38 | -0.45 | 0.07 | -0.07 | 0.54 | -0.05 | 0.69 |
| TV A velocity, m/s | 0.05 | 0.86 | -0.55 | 0.035 |  |  |  |  |
| TV E/A ratio | -0.32 | 0.45 | 0.22 | 0.59 |  |  |  |  |
| TDI MVe`, m/s | 0.11 | 0.58 | 0.06 | 0.77 | -0.32 | 0.004 | -0.15 | 0.17 |
| TDI TVe`, m/s | -0.21 | 0.42 | -0.48 | 0.05 | -0.01 | 0.93 | 0.03 | 0.77 |
| TDI MV s`, m/s | 0.14 | 0.49 | 0.14 | 0.49 | 0.04 | 0.73 | -0.02 | 0.85 |
| TDI TV s`, m/s | -0.04 | 0.89 | -0.27 | 0.30 | -0.07 | 0.55 | -0.12 | 0.29 |
| EF Biplane, % |  |  |  |  | 0.06 | 0.63 | 0.12 | 0.29 |
| GLS, % | 0.11 | 0.83 | 0.07 | 0.89 | 0.08 | 0.50 | 0.01 | 0.90 |
| RV IVRT, ms | -0.27 | 0.39 | -0.06 | 0.85 |  |  |  |  |
| LV IVRT, ms | -0.19 | 0.50 | -0.25 | 0.37 |  |  |  |  |
| E/e` | -0.25 | 0.21 | -0.42 | 0.035 | 0.24 | 0.03 | 0.11 | 0.35 |
| AV valve E velocity, m/s | -0.25 | 0.17 | -0.39 | 0.035 |  |  |  |  |
| AV valve A velocity m/s | 0.21 | 0.27 | -0.09 | 0.63 |  |  |  |  |
| AV valve E/A ratio | -0.37 | 0.045 | -0.16 | 0.41 |  |  |  |  |
| QRS duration, ms | -0.23 | 0.17 | -0.23 | 0.17 |  |  |  |  |
| VO2 max, % predicted | -0.52 | 0.23 | 0.06 | 0.89 |  |  |  |  |

IVPD, intraventricular pressure difference; MAPSE, mitral annular plane systolic excursion;
TAPSE, tricuspid annular plane systolic excursion; RV FAC, right ventricular fractional area
change; MV, mitral valve; TV, tricuspid valve; TDI, tissue doppler imaging; EF, ejection
fraction; GLS, global longitudinal strain; RV, right ventricle; LV, left ventricle; IVRT,
isovolumetric relaxation time; AV, atrioventricular; VO2 max, maximal oxygen consumption.

### Effect of respiration on IVPD based on BST

Apical 4CH BST images with respiratory data were acquired in 14 controls. The average number of acquisitions in each control were 6.5, range 4-10, and all had acquisitions during both inspiration and expiration. Total IVPD was on average 0.14 mmHg (0.55%) higher during inspiration compared to expiration (p=0.71) while apical IVPD was -0.004 mmHg (0.42%) lower during inspiration compared to expiration (p=0.89).

### Reproducibility of IVPD based on BST

The ICC (95% confidence interval) was 0.99 (0.97-0.99) for total IVPD and 0.96 (0.890-0.99) for apical IVPD for interobserver analysis. For intraobserver analysis, the ICC was 0.98 (0.86- 0.99) for total IVPD and 0.99 (0.94-0.99) for apical IVPD respectively.

### IVPD in UVH and controls

The BST analysis demonstrated that maximum negative apical pressure preceded maximal negative basal pressure (Figure 2C), and maximal total IVPD preceded peak LV inflow (Figure 2D). Median total IVPD from BST for the 30° sector in the controls was -2.22 (-1.79- -2.60) mmHg while mean apical IVPD was -0.83 ± 0.26 mmHg. There was no difference in total or apical IVPD between boys and girls (p=0.12 and 0.053 respectively).

Both total and apical IVPD increased with increasing age in the controls during the first 6 months of life. After 6 months of age, total IVPD in the controls increased with an average of -0.028 mmHg per year of age (p=0.002, correlation -0.37) (Figure 5A) while there was no significant change in apical IVPD with increasing age (Figure 5B).

There were no UVH patients < 6 months of age, and no UVH patient had apical 4CH BST acquisition with sector width < 60°. Therefore, controls < 6 months of age were excluded and IVPD from the 60° sector width in the controls was used for group comparisons. Both total and apical IVPD were significantly higher in controls compared to UVH (-1.82 vs -0.88 mmHg and -0.63 vs -0.33 mmHg, p < 0.001) (Figure 6). The difference in IVPD between the groups remained statistically significant when adjusting for differences in FR, age, heart rate (HR) and FR, age and HR combined between the groups (P < 0.001).

**Figure 6.**
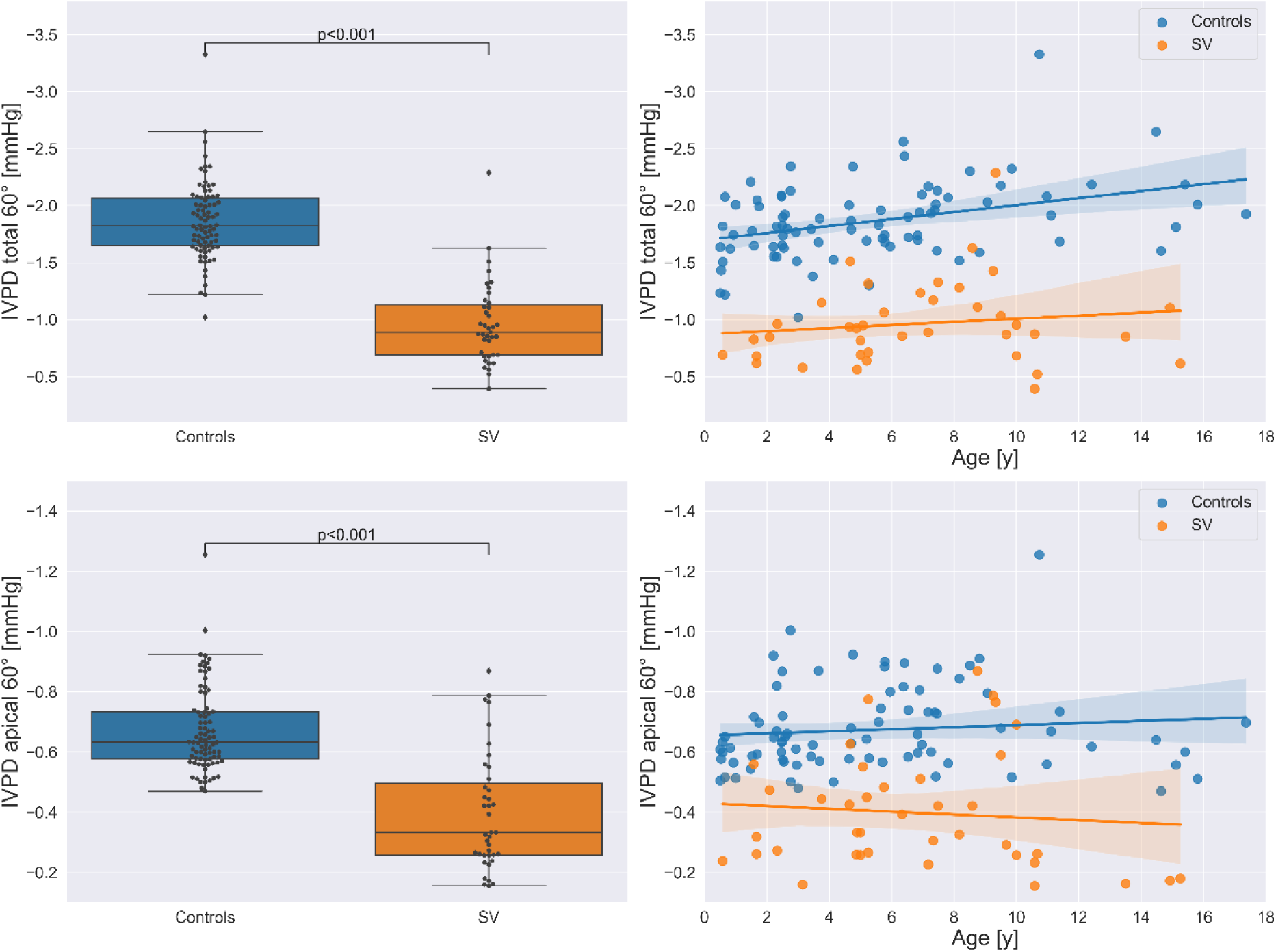
Intraventricular pressure difference (IVPD) in controls and patients with univentricular hearts (UVH) 0.5-18 years of age. Left panels: Box plots demonstrating significantly reduced total and apical IVPD in patients with UVH compared to controls. Right panels: Variation of total and apical IVPD with age including the corresponding regression line and its 95% confidence interval (shadowed area)

IVPD did not correlate with age in UVH. There was no statistical difference in IVPD between morphological RVs and LVs (p-value = 0.928 for total and 0.642 for apical IVPD respectively) or surgical stage, TCPC vs BCPC (p-value = 0.265 for total and 0.648 for apical IVPD, respectively) (Table 1).

### Clinical and echocardiographic parameters in UVH compared to controls

Comparing traditional echocardiographic parameters of diastolic function, mitral annulus early diastolic velocity by pulsed wave tissue Doppler (TDI MVè) and tricuspid annulus early diastolic velocity by pulsed wave tissue Doppler (TDI TVè) were significantly lower (p<0.001) and E/è significantly higher (p = 0.002) in UVH compared to controls.

In the controls, MV E/A ratio (correlation -0.26, P=0.022), TDI MVè (correlation -0.32, P=0.004) and E/è (correlation 0.24, p=0.03) correlated weakly but significantly with total IVPD. In UVH, AV valve E/A ratio (correlation -0.37, p=0.045) correlated weakly but significantly with total IVPD and E/è correlated significantly with apical IVPD (correlation - 0.42, p=0.035).

Looking at echocardiographic parameters of systolic function, total IVPD correlated significantly with mitral annular plane systolic excursion (correlation -0.36, p<0.001) in the controls. There was no correlation between echocardiographic parameters of systolic function and IVPD in UVH.

There were 30 UVH in New York Heart Association (NYHA) class 1 and 9 UVH in NYHA class 2. Total IVPD was -0.23mmHg lower and apical IVPD was -0.10 mmHg lower in UVH patients with NYHA class 2 compared to NYHA class 1 (p=0.094 and 0.184 respectively).

## Discussion

This study evaluates the use of early-diastolic IVPD based on BST echocardiography as a novel tool for assessment of diastolic function in children with CHD. We first validated the BST-based IVPD estimation in an invasive porcine model demonstrating that BST provides accurate estimates of early-diastolic IVPD. In a clinical validation study in healthy children, the method was found to be highly feasible and reproducible. Accuracy of BST-based IVPD requires duplex FR > 60-70 while spontaneous respiration does not have a clinically significant impact on IVPD measurements. BST-based IVPD was significantly lower in children undergoing staged UVH palliation compared to controls. There were no significant differences in IVPD when comparing pediatric UVH patients with a dominant systemic RV versus a dominant LV.

### Assessment of early relaxation and diastolic suction

Assessment of diastolic function has two main components: Ventricular relaxation and chamber stiffness.^7^ Ventricular relaxation is an active, energy-intense process. Recycling of calcium into the sarcoplasmic reticulum results in releasing the actin-myosin cross-bridges with a rapid drop in intraventricular pressure. As the repolarization wave moves from the apex to the base of the heart, the apex relaxes before the base resulting in a small intraventricular pressure difference that is present at the time of mitral valve opening and continues to rise during the first part of early filling.^24^ This pressure gradient creates a suction effect which is an important determinant of early diastolic filling and normal diastolic function.^25^ As the ventricle fills, diastolic pressure increases with end-diastolic pressure representing the diastolic pressure at maximum filling of the heart. In stiffer ventricles, filling pressures are typically elevated and rise more rapidly representing decreased compliance.

TDI MVè is a well-validated echocardiographic method to assess early relaxation in adults.^26^ When early relaxation is prolonged, e’-velocities are lower and remain low when filling pressures increase. This makes e’-velocities relatively preload-independent in the presence of a relaxation abnormality. While this concept is well established in adults, where relaxation abnormalities typically occur in the disease process before increase in filling pressures, the utility of tissue Doppler in children to detect early relaxation abnormalities is limited by the extensive variability of e’-velocities related to age, HR, heart size and filling pressures. Moreover, in children increased filling pressures can be present with preserved e’- velocities as the pathophysiology of diastolic dysfunction can be different. Alternative techniques have been explored in children, and some studies have suggested the possible applicability of early diastolic strain rate to study early relaxation parameters.^27^ As early relaxation occurs very fast, current speckle-tracking-based methods may not have the necessary temporal resolution to resolve peak early diastolic strain rate velocities.

Non-invasive echocardiographic estimation of IVPD has been proposed before based on color M-mode (CMM) and Vector flow mapping (VFM). CMM was invasively validated and has been applied clinically.^28, 29^ The one-dimensional and static nature of the method in combination with the limited anatomical and flow information available in the CMM acquisition limits the applicability of CMM-based IVPD in various cardiac conditions.^30^ VFM provides 2D IVPD based on Color Doppler imaging for radial velocities and myocardial wall speckle tracking for lateral velocities^31, 32^ This method has limitations when the radial velocity is not dominant, and the lateral velocity estimation in diastole is unreliable.^33^ In addition, the low VFM FR of around 35 frames per second (fps) results in significant underestimation of IVPD.^34^ The ability of BST echocardiography to determine IVPD based on analysis tailored to the flow pattern of each ventricle exclusively from blood flow data and with sufficient FR might offer advantages compared to CMM and VFM, especially in cases where surgical patches, variations in pressure and volume loading and anatomic variations affect blood flow patterns and wall motion.

### IVPD and diastolic function in UVH

Diastolic dysfunction in UVH patients can result from multiple factors which influence the different components of normal diastolic function. Early relaxation can be adversely affected by chronic volume and/or pressure loading with a resulting hypertrophic response and chronic hypoxemia that is typically present before the Fontan circulation.^23^ Diastolic wall motion abnormalities have been described, especially early after the Fontan operation, which can further influence early filling.^35, 36^ Diastolic dysfunction can be progressive over time as the volume-unloaded ventricle stiffens after the Fontan operation. Diastolic dysfunction significantly impacts Fontan hemodynamics as increased filling pressures reduce cardiac output related to decreased pulmonary blood flow. Moreover, it has been suggested that pulmonary blood flow in UVH is highly dependent on early diastolic filling properties of the ventricle.^37^ It is, however, extremely difficult to detect early relaxation abnormalities in UVH patients related to intrinsic difficulties with AV valve inflow velocities and tissue Doppler velocities. Diastolic suction can be affected by both decreased early relaxation and incoordinate wall motion and could be a useful parameter for assessing early diastolic function. Our finding of significantly reduced IVPD in early diastole in patients with UVH compared to controls indicates impaired early relaxation and reduced diastolic suction. We hypothesize this could be an important early marker of diastolic ventricular dysfunction that is difficult to detect using the conventional parameters.

IVPD is found to be stable despite considerable preload reduction including during mitral occlusion producing non-filling diastole.^38, 39^ Interestingly we found no difference between the patients after TCPC versus BCPC. This seems to confirm this is a relative preload independent measurement as a significant preload reduction occurs after Fontan completion. Based on this, we do not consider it likely that any difference in preload between UVH and controls could explain the finding of reduced IVPD in UVH in our study.

We observed that tissue Doppler velocities are lower in UVH patients versus healthy controls, but this could be an effect of loading conditions. The prognostic value will need further validation.

We observed a similar reduction in IVPD in dominant RV and LV variants of UVH, suggesting that the effect on early relaxation and diastolic suction occurs in both morphologies. This could potentially be explained by the RV remodeling process in which UVH with morphological RVs adopt an LV functional pattern. ^40^

### Total or regional IVPD

Studies have suggested that IVPD from the mid-to-apical part of the LV is a more accurate measure of relaxation and more sensitive than total or basal IVPD in diagnosing diastolic dysfunction ^41, 42, 43, 44^ In adult patients, basal IVPD is typically defined as IVPD from the basal 2 cm of the LV and apical IVPD as IVPD from the remaining apical part of the LV. Studies on pediatric patients have defined basal IVPD as IVPD from the basal 1/3 of the LV and apical IVPD as IVPD from the apical 2/3 of the LV to account for variation in LV size in the pediatric population.^43, 45^ There is no established consensus on how to make the distinction between IVPD from the basal and apical part of the ventricle, and in the present study we have adopted the division used in previous pediatric studies.

Invasive measurements have demonstrated the presence of a continuous pressure gradient with gradual increase in minimal LV pressure from apex to base indicating that all regions of the LV are capable of creating early-diastolic suction.^8^ Our data demonstrate significant differences between UVH and controls for both total, basal and apical IVPD (Table 1) and do not provide evidence to conclude on whether total or regional IVPD is preferable for assessment of diastolic function in children with UVH.

### IVPD in pediatric and adult cardiology

We have previously demonstrated significantly reduced early diastolic IVPD using BST in children with hypertrophic and dilated cardiomyopathies.^19^ These are patient groups expected to have diastolic dysfunction where current echocardiographic methodology has limitations for the assessment of diastolic function.^4^ Clinical studies both in children and adults have demonstrated the potential of IVPD in early diastole as a sensitive and early predictor of diastolic dysfunction in a variety of cardiac conditions. ^43,44,45,46^ Diagnosing HFpEF using IVPD could be of particular value in the adult population.^41,32^ More recently IVPD analysis based on BST has become feasible in adults.^24^

### Optimizing BST acquisitions for IVPD analysis

In vitro and in silico data indicate that FR > 60 are required for accurate IVPD- estimates.^34, 47^ The BST acquisitions with 30 dg sector width in the 118 controls in our study had a median duplex FR of 94 (range 64-161) fps. In the invasive validation part of this study, median duplex FR was 79 (68**-**82) fps. These data indicate that BST acquisitions can achieve sufficient FR for accurate IVPD-estimates in the clinical setting.

Spontaneous respiration did not significantly affect IVPD in the 14 controls with respiration accounted for in the present study. In healthy hearts, MV peak inflow velocity has a maximum respiratory variation of 15%.^48^ Given end-systolic volume does not seem to be significantly affected by respiration,^49, 50^ it seems plausible that relaxation properties of the myocardium is not affected by respiration and that respiratory AV-valve inflow velocity variation is caused by other factors.

### Limitations

The invasive validation was performed in an open chest porcine model with the probe directly on the heart, resulting in echocardiographic image quality often not achievable in clinical cardiology. Enrollment of patients with UVH and controls was done at different centers. IVPD-results based on BST is dependent on image quality and clutter filtering, and technical settings must be standardized as in this study. High frame rate plane waves have limited depth penetrance, and velocity estimates based on BST using the 6S pediatric probe are accurate to depths of 8-10 cm.

BST applies 2D technology on three-dimensional (3D) flow. The validity of this could be questioned. A 3D computer flow model demonstrated validity of computing IVPD from 2D velocities.^19^ In the invasive study presented here, the pressure catheters were often not visible on the BST images used for IVPD analysis, so invasive pressure measurements and BST-based pressure estimates could be from different planes, yet they demonstrated excellent agreement. Reproducibility of IVPD was excellent when comparing multiple standardized BST acquisitions in the same patient from two different cardiologists in the present study. These data suggest validity of using 2D BST imaging in the apical 4CH plane for estimating IVPD during early diastolic filling.

## Conclusions

BST echocardiography allows accurate non-invasive estimation of IVPD, a measure of ventricular early diastolic relaxation and suction. IVPD is significantly reduced in children with UVH compared to controls indicating impaired relaxation in patients with UVH. IVPD based on BST has potential to improve assessment of diastolic function in a variety of cardiac conditions both in children and adults, however, the clinical and prognostic value of the method and technology requires large clinical studies.

## Data Availability

All echocardiographic basic and raw data and all clinical data referred to in the manuscript are available for investigation on request.

## Non-standard Abbreviations and Acronyms

2D: Two dimensions, two-dimensional
3D: Three-dimensional
4CH: Four-chamber
AV: Atrioventricular
BCPC: Bidirectional cavopulmonary connection
BST: Blood speckle tracking
CHD: Congenital heart disease
CMM: Color M-Mode
FPS: Frames per second
FR: Frame rate
HR: Heart rate
ICC: Intraclass correlation coefficient
IVPD: Intraventricular pressure difference
LV: Left ventricle, left ventricular
MV: Mitral valve
NYHA: New York Heart Association
RV: Right ventricle
TCPC: Total cavopulmonary connection
TDI MVè: Mitral annulus early diastolic velocity by pulsed wave tissue doppler
TDI TVè: Tricuspid annulus early diastolic velocity by pulsed wave tissue doppler
UVH: Univentricular hearts
VFM: Vector Flow Mapping

## Acknowledgements

We thank Wei Hui and Cameron Slorach for data collection at the Hospital for Sick Children.

## Supplemental materials

IVPD from BST, mathematical formula and method.

## Supplemental data

Intraventricular pressure differences (IVPD) from BST: The pressure gradients are estimated from the velocity field as follows ∂ *P* ∂ *x* = −*ρ* ( ∂ *v x* ∂ *t* + *v x* ∂ *v x* ∂ *x* + *v y* ∂ *v x* ∂ *y*) + *μ* ∇ 2*v x* , Eq. 1 ∂ *P* ∂ *y* = −*ρ* ( ∂ *v y* ∂ *t* + *v y* ∂ *v y* ∂ *y* + *v x* ∂ *v y* ∂ *x*) + *μ* ∇ 2*v y* , Eq. 2 where ρ = 1060 kg/m3 is the density of the blood, v = [vx, vy] is the blood velocity vector and µ = 0.004 Pa·s is the blood viscosity. IVPD is found by integrating the pressure gradients along a manually defined path in the LV from base to apex. *I V P D* = ∫ ∂ *P* ∂ *x d x* + ∫ ∂ *P* ∂ *y d y* Eq. 3 The path is a spline curve defined by three or more points set by the user according to the mitral inflow streamlines. Positive IVPD values depict acceleration of blood towards the base or deceleration of flow which originally was directed towards the apex. Negative IVPD values depict acceleration of blood towards the apex or deceleration of flow originally directed towards the base. The unit of IVPD is millimeters of mercury (mmHg)

